# Persistence of immune response in health care workers after two doses BNT162b2 in a longitudinal observational study

**DOI:** 10.1101/2021.12.17.21267926

**Authors:** Jonas Herzberg, Bastian Fischer, Christopher Lindenkamp, Heiko Becher, Ann-Kristin Becker, Human Honarpisheh, Salman Yousuf Guraya, Tim Strate, Cornelius Knabbe

**Affiliations:** Department of Surgery – Krankenhaus Reinbek St. Adolf-Stift, Hamburger Strasse 41, 21465 Reinbek, Germany; Institut für Laboratoriums- und Transfusionsmedizin, Herz- und Diabeteszentrum NRW, Georgstraße 11, 32545 Bad Oeynhausen, Germany; Institute of Medical Biometry and Epidemiology, University Medical Center Hamburg-Eppendorf, Martinistrasse 52, 20246 Hamburg, Germany; Asklepios Klinik Harburg, Abteilung für Psychiatrie und Psychotherapie, Eißendorfer Pferdeweg 52, 21075 Hamburg, Germany; Clinical Sciences Department, College of Medicine, University of Sharjah, P. O. Box 27272 Sharjah, United Arab Emirates

**Keywords:** SARS-CoV-2, humoral and cellular immunity, Health Care worker, immunological memory, vaccination, BNT162b2

## Abstract

**Background:** The mRNA-based vaccine BNT162b2 of BioNTech/Pfizer has shown high efficacy against SARS-CoV-2 infection and a severe course of the COVID-19 disease. However, little is known about the long-term durability of the induced immune response resulting from the vaccination.

**Methods:** In a longitudinal observational study in employees at a German hospital we compared the humoral and cellular immune response in 184 participants after two doses of the BioNTech/Pfizer vaccine (BNT162b2) with a mid-term follow-up after 9 months. Anti-SARS-CoV-2 binding antibodies were determined using both a quantitative and a semi-quantitative assay. For a qualitative assessment of the humoral immune response, we additionally measured neutralizing antibodies. Cellular immune response was evaluated by measuring Interferon-gamma release after stimulating blood-cells with SARS-CoV-2 specific peptides using a commercial assay.

**Results:** In the first analysis, a 100% humoral response rate was described after two doses of BNT162b2 vaccine with a mean antibody ratio of 8.01 ± 1.00. 9 months after the second dose of BNT162b2, serological testing showed a significant decreased mean antibody ratio of 3.84 ± 1.69 (p < 0.001). Neutralizing antibodies were still detectable in 96% of all participants, showing an average binding inhibition value of 68.20% ± 18.87%. Older age, (p < 0.001) and obesity (p = 0.01) had a negative effect on the antibody persistence. SARS-CoV-2 specific cellular immune response was proven in 75% of individuals (mean IFN-gamma release: 579.68 mlU/ml ± 705.56 mlU/ml).

**Conclusion:** Our data shows a declining immune response 9 months after the second dose of BNT162b2, supporting the potentially beneficial effect of booster vaccinations, the negative effect of obesity and age stresses the need of booster doses especially in these groups.

## Introduction

In order to fight the global coronavirus disease 2019 (COVID-19) pandemic, a variety of vaccines were applied worldwide, beginning in December 2020, and were felt to be the pivoting point in this pandemic situation. The mRNA vaccines, provided by BioNTech/Pfizer (BNT162b2) and Moderna (Spikevax) have shown high efficacy in clinical trials and real-world-data [1,2]. Until now, real-world-data regarding the persistence of the induced humoral and especially cellular immune response post-vaccination are rare.

Even in short-term studies, elderly people were reported to have a less intense immune response after two doses of the BioNTech/Pfizer vaccine BNT162b2 [3,4]. This was also seen in patients under immunosuppression, such as after organ transplantation [5].

Initial studies reported a decrease in antibodies after 6 months [4,6] or a correlating increased number of infections after complete vaccination [7]; but data regarding the longitudinal serological dynamics of immunization after BNT162b2 vaccine is limited.

In this trial we report the mid-term dynamics of immune response, 9 months after second dose of BNT162b2, by determining anti-SARS-CoV-2-immunglobulin G (IgG) antibodies, T-cell-response and neutralizing antibodies. The study cohort consisted of a well-defined group of health care employees, known to be under a higher risk for COVID-19.

## Methods

### Study design

In April 2020 we initiated a longitudinal study in health care workers measuring sero-epidemiological data during the COVID-19 pandemic [8]. All employees of the secondary care hospital located in the province of Schleswig-Holstein near the border of the city of Hamburg in Northern Germany were invited to participate. All employees were offered vaccination against SARS-CoV-2 starting in December 2020. In April 2021, all participants were invited to provide a blood specimen to evaluate the antibody prevalence after vaccination or infection [9].

For this study, an additional analysis of all participants around 9 months after second dose of BioNTech/Pfizer vaccine was made to evaluate the longitudinal persistence of the vaccine-induced immune response.

All blood samples were collected on November 13^th^ – 14^th^ 2021 and all participants were asked to complete an additional questionnaire, regarding post-vaccination reactions and previous SARS-CoV-2 infections.

All participants provided written and informed consent prior to enrolment. This study was prospectively registered at the German Clinical Trial Register (DRKS00021270) after approval by the Ethics Committee of the Medical Association Schleswig-Holstein. All study activities were conducted in accordance with the Declaration of Helsinki.

### Anti-SARS-CoV-2-IgG antibodies

The fully automated semiquantitative anti-SARS-CoV-2-ELISA (IgG) from Euroimmun (Lübeck, Germany) was used to detect the S1 domain of the SARS-CoV-2 spike-protein. In accordance to the manufacturer this test shows a specificity of 99.0% and sensitivity of 93.8% [10]. As this was the same test used within the previous analyses, a longitudinal comparability was ensured. A ratio below 0.8 was considered negative, a ratio ≥ 0.8 to < 1.1 was considered equivocal, and a ratio ≥ 1.1 was considered positive as defined by the manufacturer.

In addition, a fully automated quantitative anti-SARS-CoV-2-assay (IgG) from Abbott (Chicago, USA) was performed. In keeping with the WHO-standard, data were expressed in Binding Antibody Units per ml (BAU/ml). Samples were marked seronegative below 7.1 BAU/ml whereas values above 7.1 BAU/ml were determined to be positive, as mentioned by the manufacturer.

### Neutralizing antibodies against SARS-CoV-2

All samples were analyzed for neutralizing anti-SARS-CoV-2 antibodies using the NeutraLISA™ SARS-CoV-2 Neutralization Antibody Detection KIT (Euroimmun, Lübeck, Germany) in accordance to the manufacturer’s instructions. Binding inhibition values above 35% were considered positive, whereas values between 20% and 35% were considered equivocal.

### T-cell-response

Cellular immunity to SARS-CoV-2 was assessed by using an Interferon (IFN)-gamma release assay (IGRA) from Euroimmun (Quan-T-cell SARS-CoV-2 kit). The assay was performed according to manufacturer’s instructions. In brief, 500 μl of heparinized blood was stimulated with SARS-CoV-2 specific peptides covering regions of the viral S1-domain. After incubating the tubes (37°C, 22 h), plasma was collected and tested for Interferon-gamma release using an ELISA-assay (Quan T-cell ELISA, Euroimmun). Background IFN-gamma values were assessed by incubating blood without prior peptide-stimulation. As a positive control, blood cells were stimulated with mitogen, resulting in a broad and unspecific IFN-gamma secretion. IFN-gamma concentration was expressed as mIU/ml.

### Statistical analysis

IBM SPSS Statistics Version 25 (IBM Co., Armonk, NY, USA) was used for statistical analysis. Graphics were elaborated using IBM SPSS Statistics Version 25 (IBM Co., Armonk, NY, USA) and GraphPad Prism 9.

All variables are presented as means with standard deviation. Categorical variables are shown as numbers with percentages. Fisher’s exact test or chi-square test was used to determine relationships between categorical variables depending on size of groups. Exact 95% confidence intervals were provided where appropriate. Differences between groups were analyzed using Wilcoxon test. Inter-group differences were analyzed using Mann-Whitney-U test or Kruskal-Wallis-test. A linear regression analysis was done to investigate the joint effect of age, sex, body mass index and current smoking on antibody and t-cell response using the backward selection method. The t-cell response had a skewed distribution and was logarithmized for the regression analysis. A p-value < 0.05 was considered statistically significant.

## Results

A total of 184 participants provided a blood sample 9 (range 7-9 months) months after receiving their second dose of BioNTech/Pfizer. This meant a follow-up rate of 58.41% compared to the analysis after the second dose of vaccine (315 participants in April 2021) [9].

In this follow-up, the study characteristics did not differ significant to the previous follow-ups with 73.4% female and 26.6% male and a mean age of 46.32 ± 10.91 years. 3 participants reported a previous SARS-CoV-2 infection. Of these, 1 was reported prior to vaccination, and 2 cases occurred between the second dose of vaccine and this follow-up (**Table 1**). These participants are included in the following analysis.

**Table 1:** Immune status of previously infected participants in comparison to the mean values of the study cohort.

| Participant | Time since infection [months] | Anti-SARS-CoV-2-IgG [BAU/ml] | Neutralizing antibodies [%] | IFN-gamma [mIU/ml] |
| --- | --- | --- | --- | --- |
| A | 17.5 | 314.40 | 97.26 | 645.60 |
| B | 8.5 | 304.30 | 96.87 | 170.00 |
| C | 6.5 | 26.90 | 39.12 | 1216.20 |
| **Study cohort** |  | **Anti-SARS-CoV-2-IgG [BAU/ml] (mean ± SD)** | **Neutralizing antibodies [%] (mean ± SD)** | **IFN-gamma [mIU/ml] (mean ± SD)** |
| n = 184 | ----- | 124.67 ± 104.41 | 68.20 ± 18.87 | 579.68 ± 705.56 |

### Anti-SARS-CoV-2-IgG

In the previous analysis, all participants showed a positive antibody-ratio after two doses of BioNTech/Pfizer, whereas two participants seroconverted to an equivocal result in this follow-up after 9 months. The antibody-ratio was significantly lower in the follow-up analysis after 9 months (8.01 ± 1.00. vs. 3.84 ± 1.69; p < 0.001).

The mean reduction of the IgG antibody-ratio was 53.11% ± 17.95%.

In order to further improve the assessment of the humoral immune response, quantification of anti-SARS-CoV-2 antibodies was performed according to WHO standards (BAU/ml). All participants showed antibody levels above the manufacturer’s cutoff (mean: 124.67 ± 104.41 BAU/ml) (**Figure 1**).

**Figure 1:**
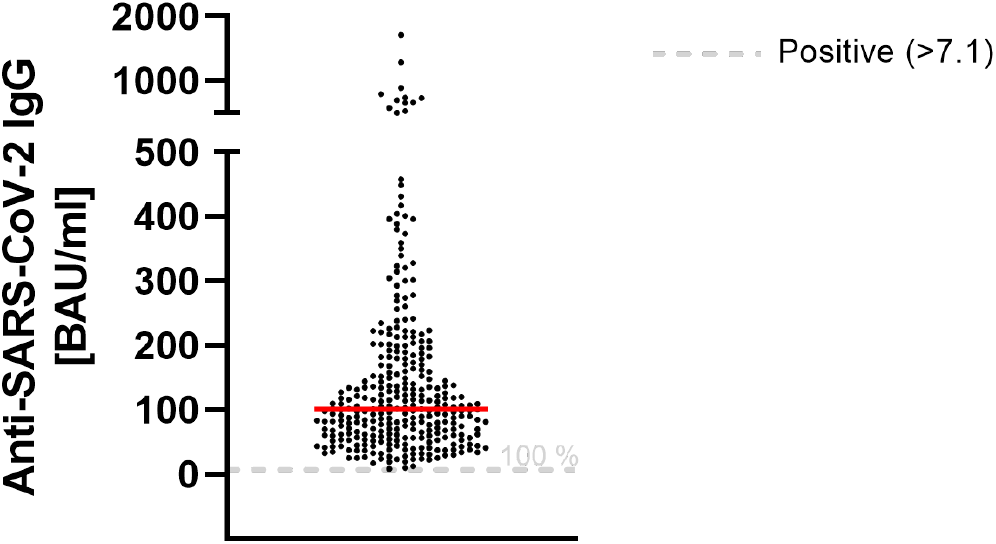
Quantitative analysis of anti-SARS-CoV-2-IgG 9 months after second dose of BNT162b2 Red line marks the mean. The dashed gray line marks the positive cutoff specified by the manufacturer.

### Neutralizing antibodies

Overall, 96% of study-participants showed neutralizing antibodies against SARS-CoV-2. Our data show a mean binding inhibition capability of 68.20% ± 18.87% 9 months after the second vaccination using BioNTech/Pfizer vaccine (**Figure 2**).

**Figure 2:**
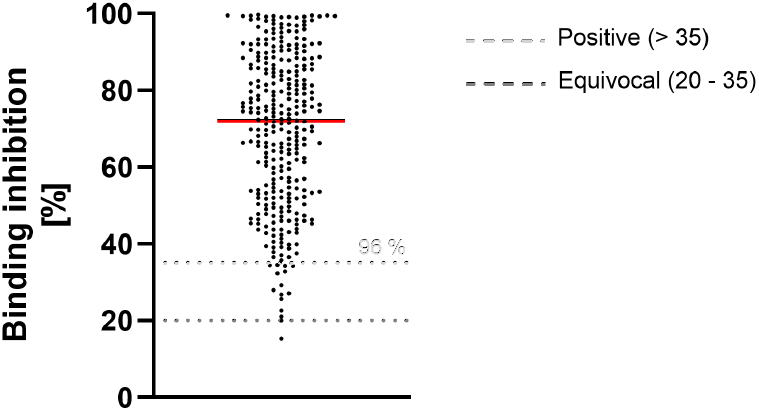
Binding inhibition of neutralizing antibodies 9 months after vaccination with BioNTech/Pfizer Red line marks the mean. The dashed lines show the positive (light gray)- and equivocal (dark gray) cutoffs specified by the manufacturer.

### T-cell response

9 months after the second dose of the BioNTech/Pfizer vaccine, 73.4 % of participants had a detectable T-cell-immune response. There was no significant correlation of the positivity in relation to the sex, obesity or smoking behavior. The mean IFN-gamma concentration was 579.68 mlU/ml ± 705.56 mlU/ml within this study cohort (**Figure 3**).

**Figure 3:**
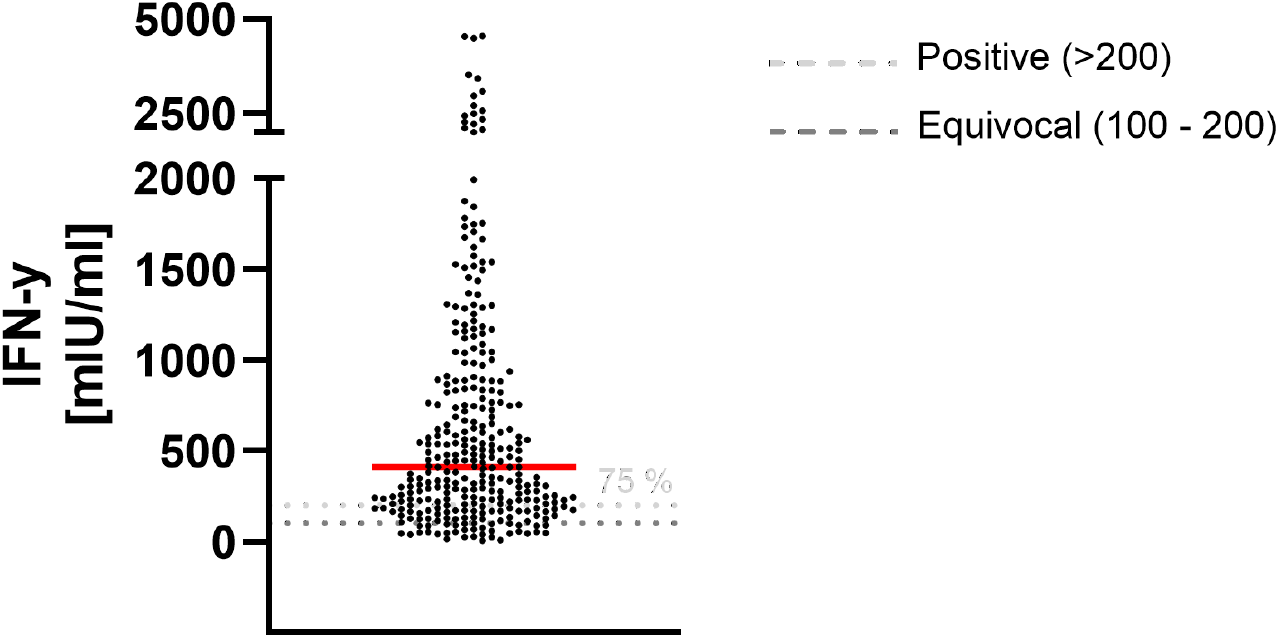
IFN-gamma measured as T-cell-response 9 months after second dose of BioNTech/Pfizer Red line marks the mean. The dashed lines show the positive (light gray)- and equivocal (dark gray) cutoffs specified by the manufacturer.

We identified several factors associated with a lower T-cell response and a lower level of neutralizing antibodies (**Table 2**). A BMI above 30 (p = 0.004) and smoking (p = 0.034) was associated with a reduced level of neutralizing antibodies. Male participants showed a lower T-cell response at this timepoint, whereas this difference remained insignificant following the Mann-Whitney-U-test.

**Table 2.** Participants 9 months after second dose of BioNTech/Pfizer vaccine (n=184) M: mean SD: standard deviation ^a^ Mann-Whitney-U-test ^b^ Kruskal-Wallis-Test Obesity is defined as a Body mass index above 30.

|  | Antibody ratio after 9 months, M ±SD | p-value | Reduction of initial antibody response (%), M ±SD | p-value | T-cell response, M ±SD | p-value | Neutralizing antibodies (%), M ±SD | p-value |
| --- | --- | --- | --- | --- | --- | --- | --- | --- |
| All (n=184) | 3.84 ±1.69 |  | 53.11 ± 17.95 |  | 579.68 ± 705.56 |  | 68.20 ± 18.87 |  |
| Sex |  |  |  |  |  |  |  |  |
| Male (n=49) | 3.84 ± 1.71 | 0.938<sup>a</sup> | 52.32 ± 17.98 | 0.712<sup>a</sup> | 492.17 ±528.48 | 0.146<sup>a</sup> | 66.98 ± 20.93 | 0.805<sup>a</sup> |
| Female (n=135) | 3.84 ± 1.68 |  | 53.39 ± 18.00 |  | 611.44 ± 758.94 |  | 68.65 ± 18.12 |  |
| Smoking |  |  |  |  |  |  |  |  |
| Yes (n= 52) | 3.45 ± 1.56 | 0.062<sup>a</sup> | 56.80 ± 16.25 | 0.120<sup>a</sup> | 512.60 ± 726.06 | 0.162<sup>a</sup> | 63.59 ± 18.84 | 0.034<sup>a</sup> |
| No (n= 132) | 3.99 ± 1.72 |  | 51.65 ± 18.43 |  | 606.10 ± 698.35 |  | 70.03 ± 18.64 |  |
| Obesity |  |  |  |  |  |  |  |  |
| Yes (n= 30) | 3.19 ± 1.70 | 0.012<sup>a</sup> | 59.16 ± 18.82 | 0.020<sup>a</sup> | 451.80 ± 475.61 | 0.306<sup>a</sup> | 58.68 ± 20.20 | 0.004<sup>a</sup> |
| No (n= 154) | 3.96 ±1.66 |  | 51.92 ± 17.60 |  | 604.58 ± 740.75 |  | 70.07 ± 18.09 |  |
| Age groups |  |  |  |  |  |  |  |  |
| <30 (n= 13) | 4.92 ± 1.95 | 0.003<sup>b</sup> | 42.86 ± 21.82 | 0.008<sup>b</sup> | 465.95 ± 523.68 | 0.216<sup>b</sup> | 78.60 ± 19.89 | 0.001<sup>b</sup> |
| 31-39 (n= 40) | 4.45 ± 1.66 |  | 46.99 ± 18.08 |  | 753.12 ± 980.72 |  | 74.37 ± 18.62 |  |
| 40-49 (n= 57) | $3.77 \pm 1.47$ | | $53.58 \pm 16.17$ | | $568.40 \pm 678.75$ | | $69.32 \pm 15.95$ | |
| 50-59 (n= 47) | $3.36 \pm 1.61$ | | $57.65 \pm 16.80$ | | $468.27 \pm 565.18$ | | $62.07 \pm 20.29$ | |
| >60 (n= 27) | $3.36 \pm 1.75$ | | $58.19 \pm 17.89$ | | $595.19 \pm 549.00$ | | $62.13 \pm 17.48$ | |

During the 9 months follow-up period, the antibody-ratio showed a significant decrease in older participants (p = 0.003 following the Kruskal-Wallis-test) (**Figure 4**).

**Figure 4:**
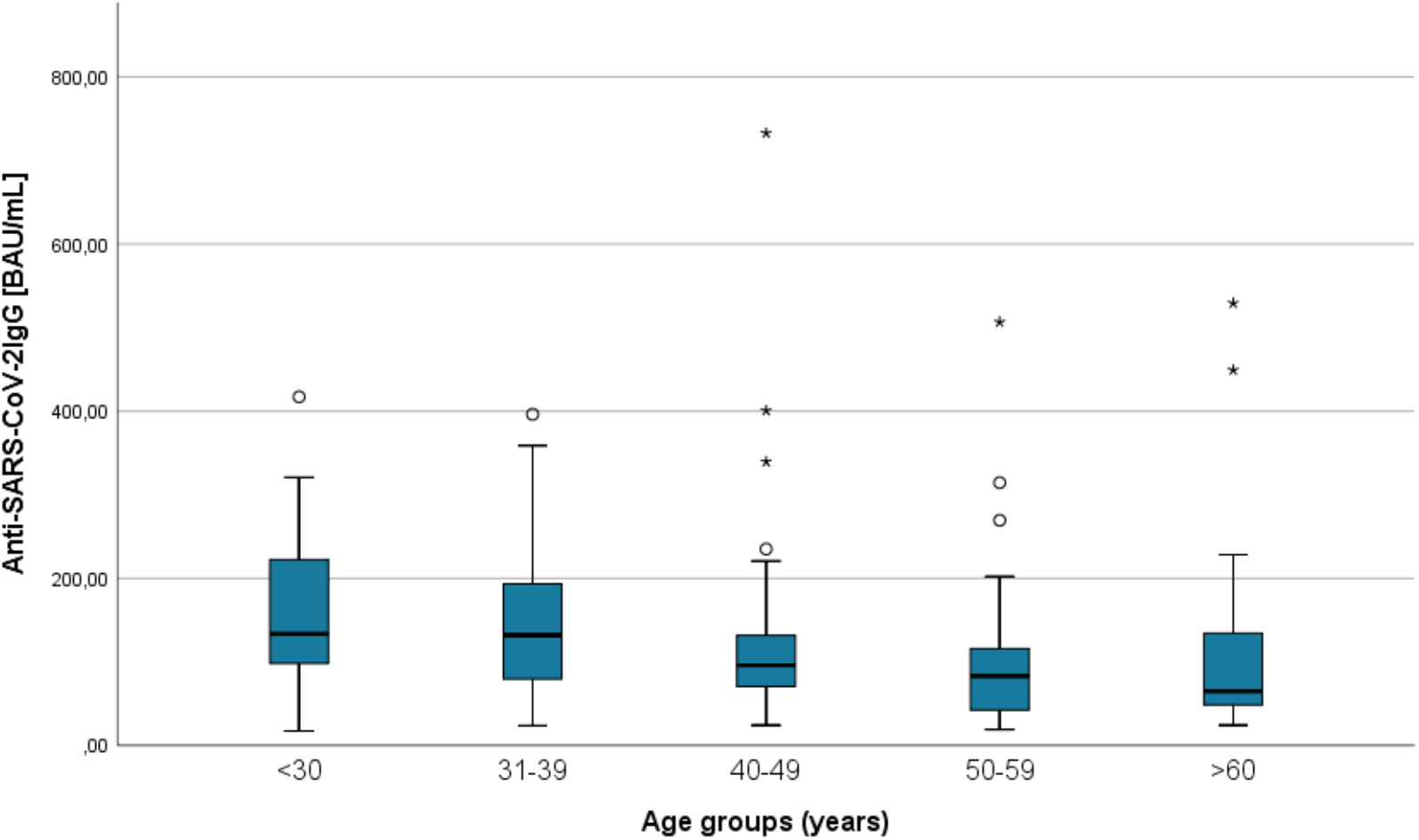
Age dependent anti-SARS-CoV-2-IgG 9 months after second dose of BioNTech/Pfizer vaccine with a significantly reduced ratio in older participants (p=0.003)

There was also a statistically significant decrease in neutralizing antibodies (p = 0.001) (**Figure 5**), but the reduction in INF-gamma-level remained statistically insignificant (p = 0.218).

**Figure 5:**
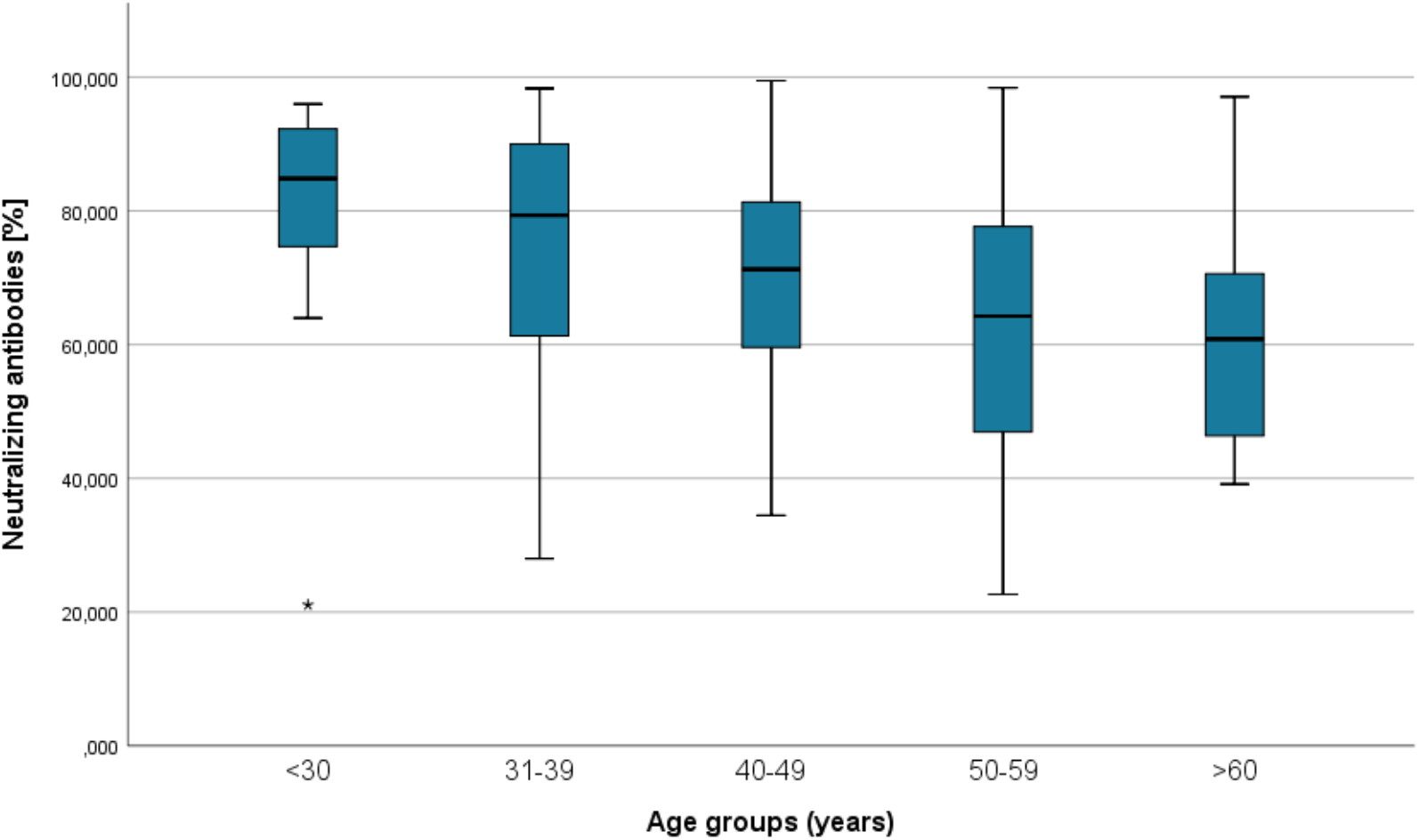
Age dependent neutralizing antibodies 9 months after second dose of BoNTech/Pfizer showing a significant reduction in older participants (*p* = 0.001).

The additionally performed linear regression analysis confirmed the univariate analyses regarding the differences in persisting immune response after 9 months. A significant negative effect on antibody persistence was observed for older (p<0.001) and obese (p=0.01) participants. Smoking had also a negative effect, with p-value slightly above the limit (p=0.08) **Table 3** gives the regression estimates and the corresponding p-values and 95% confidence intervals. Sex had no effect. The same result was also found for neutralizing antibodies as dependent variable. **Table 4** gives the corresponding results. Thus, the estimated neutralizing antibodies value for a nonsmoker, age 50 and BMI 25 is 68.705%.

**Table 3:** Linear regression for Anti-SARS-CoV-2 antibody ratio as dependent variable and smoking, age and BMI as covariable.

| Variable |  | Parameter Estimate | p-value | 95% CI |
| --- | --- | --- | --- | --- |
| (Intercept) | $\alpha$ | 1.249 | <0.001 | (1.168, 1.329) |
| BMI -25* | $\beta_1$ | -0.0182 | 0.01 | (-0.032,- 0.0043) |
| Current smoking (yes) | $\beta_2$ | -0.128 | 0.08 | (-0.273, 0.017) |
| Age -50* | $\beta_3$ | -0.0115 | <0.001 | (-0.0175,- 0.00544) |

CI: confidence interval; BMI: body mass index
$R^2=0.13$
\*linear transformation of BMI (minus 25) and Age (minus 50)

**Table 4:** Linear regression for neutralizing antibodies as dependent variable and smoking, age and BMI as covariable.

| Variable |  | Parameter Estimate | p-value | 95% CI |
| --- | --- | --- | --- | --- |
| (Intercept) | $\alpha$ | 68.705 | <0.001 | (65.47, 71.94) |
| BMI -25* | $\beta_1$ | -0.687 | 0.02 | (-1.24, -0.13) |
| Current smoking (yes) | $\beta_2$ | -5.51 | 0.06 | (-11.32, 0.303) |
| Age -50* | $\beta_3$ | -0.423 | <0.001 | (-0.665, -0.180) |

CI: confidence interval; BMI: body mass index
$R^2=0.13$
\*linear transformation of BMI (minus 25) and Age (minus 50)

## Discussion

This study analyzes the persistence of the humoral and cellular response, including neutralizing antibodies, to BioNTech/Pfizer vaccine in a well-defined group of hospital employees as part of a longitudinal evaluation.

Up until now, little is known about the persistence of the immune response after vaccination with BioNTech/Pfizer. This study reveals a significant antibody decrease in a mid-term-follow-up in all subgroups, affecting especially older people and people with a high BMI.

### Immune response after 9 months

Currently there is minimal data about the long-lasting effect following active immunization with mRNA vaccines.

In our cohort, the antibody ratio decreased in the 9-month follow-up to about the half of the initial value. This was not surprising, as first half-year follow-ups showed a decrease during that timeframe [4], and it is well known, that not all plasmablasts commit as memory plasma cells [11,12].

The decreased antibody ratio is also detected in the decreased neutralization capacity. Several studies found this decrease to begin as early as 3 months after vaccination [4,13]. Discussing the long-lasting effect of mRNA vaccines (such as the one from BioNTech/Pfizer) in protecting against COVID-19 disease, the role of cellular immunity in addition to humoral response might be important but is often disregarded due to elaborate assays, which are not feasible in standard-laboratories. We determined cellular immunity in a simplified approach by IFN-gamma release after stimulation of blood-cells with specific SARS-CoV-2 peptides.

In our cohort, 73.40% showed a detectable T-cell response even 9 months after vaccination. This is in line with data presented by Naaber et al. for their 6-months follow-up [4], and in keeping with the initial phase I/II trial showing an activation of T-cells after using mRNA vaccines [14].

Compared to our data, Tober-Lau et al. determined a higher median IFN-gamma release six months after BNT162b2-vaccination within their HCW-cohort using the same IGRA-assay (1198.0 mIU/ml vs. 412.0 mIU/ml) [15]. In addition, binding inhibition capability of neutralizing antibodies was considerably lower within our cohort (88.1 % vs. 68.2 %). Reasons for this may be the different time points of examination after second vaccination and the higher average age of our cohort.

### Risk factors for a reduced immune response to SARS-CoV-2

In our follow-up study, we found a correlation between age, obesity and smoking with respect to almost all considered immune responses.

As shown in previous studies, we confirmed the negative correlation between antibody responses and the age of vaccinated individuals [3,4].

In addition to the reduced antibody response after vaccination older people are known to have a faster decrease after vaccination [4].

Interestingly, obese participants with a BMI higher than 30 showed a significantly higher decrease in the antibody ratio and also significantly lower neutralizing antibodies not only in the univariate analysis but also following the regression analysis. This effect was first described by Watanabe et al. [16] and seen by Malavazos et al. [17] in the first months after vaccination.

The effect of smoking immediately after vaccination was already discussed [9,16] and is reported to influence the effectiveness of other vaccines due to a general immunosuppression caused by smoking [16,18]. In this 9-month evaluation, for the only significant difference was with respect to neutralizing antibodies.

These findings are important as obese people and elderly are known to have a higher risk not only for SARS-CoV-2 infection, but especially for a severe course of COVID-19 [19,20].

Further studies are needed to evaluate the longitudinal course of cellular and humoral immune response not only after two doses of BioNTech/Pfizer but also after a third dose.

### Limitation

The major limitation of this trial is its single-center design. Due to the inclusion of hospital employees, women are relatively overrepresented and other groups with a higher risk are underrepresented. Especially elderly participants, with an age over 70 years, are not included in this study.

It cannot be excluded that there were asymptomatic, undetected SARS-CoV-2 infections among the participants during the 9 months after second vaccination, which may lead to a slight bias in the results.

Due to the use of different methods, it was not possible to compare the absolute values of antibody concentrations, T-cell responses or neutralizing antibodies over the follow-up-period. Further evaluations of antibody response after vaccination are needed, to investigate the longitudinal persistence of antibodies and the possible need for further booster vaccinations.

## Conclusion

This study showed an age-dependent decrease of vaccine-induced anti-SARS-CoV-2-IgG antibodies in a midterm-follow-up after 9 months. People with a higher risk for a severe COVID-19-disease course, such as elderly or obese people, showed a reduced immune response at this timepoint after vaccination. These results encourage the use, and need, of so-called booster doses 6 months after the second dose of the BioNTech/Pfizer vaccine BNT162b2, especially in vulnerable individuals such as elderly or obese people.

## Data Availability

All data produced in the present study are available upon reasonable request to the authors.

## Acknowledgments

We would like to thank all team members at the Krankenhaus Reinbek St. Adolf-Stift, and all participating laboratories that analyzed these additional samples in a phase of high workload. Thanks also to Rebecca Zimmer (linguistic enrichment) for her intense and rapid help.

